# Surveillance-based estimation of the impact of introducing a pathfinder programme for nirsevimab immunisation in Ireland on infant hospitalisations due to respiratory syncytial virus in 2024/2025

**DOI:** 10.1101/2025.04.23.25326289

**Authors:** Darren L Dahly, Katie O’Brien, Lisa Domegan, Maureen O’Leary, Eva Kelly, Michael Hanrahan, Adele McKenna, Ajay Oza, Augustine Pereira, Éamonn O’Moore

## Abstract

Respiratory syncytial virus (RSV) presents a significant health risk to young infants and substantially burdens already stretched health systems every winter. Ireland launched a RSV immunisation pathfinder programme for newborns and other clinically vulnerable infants in September of 2024. While a comprehensive evaluation of the programme is underway, we aimed to rapidly analyse the impact of the programme on infant RSV hospitalisations. Based on an analysis of notified RSV hospitalisations spanning this and four previous seasons using generalised additive models, we estimated that there was a 75% reduction in the incidence rate for nirsevimab eligible infants born this season, conditional on the age-group and season effects adjusted for in the model (incidence rate ratio 0.25; 95% CI 0.19 to 0.33). This corresponded to an estimated 532 RSV hospitalisations averted (95%CI 369 to 695) for this group, in this season.

## Introduction

Respiratory syncytial virus (RSV) infection, though mild for most people, is a leading cause of hospitalisation for infants under 1 year of age, and can have serious consequences for older adults and people with cardiopulmonary disease and immunocompromising conditions (1–4). RSV has a well-established seasonal pattern, with most infections occurring in the late autumn and winter months, frequently overlapping with seasonal influenza (5) and potentially with COVID-19. This pattern places winter pressures on health systems, which can have knock-on effects on population health (6,7).

Nirsevimab is a human recombinant monoclonal antibody that can prevent RSV infection or mitigate its effects. It has been approved for use in the European Union since late 2022 (8,9), based in part on evidence of efficacy and safety from phase 3 randomised controlled trials (10). A single dose is given directly to infants, and provides passive immunity against RSV for approximately six months (11). This window is critical given that the highest burden of disease is in the neonatal (0-27 days) and early infant periods (28-183 days). Following European Medicines Agency approval, Spain launched a nirsevimab newborn immunisation programme for the 2023/2024 RSV season, and evidence suggests it substantially reduced infant RSV hospitalisations (12,13).

In April 2024, the Irish National Immunisation Advisory Committee (NIAC) recommended passive immunisation of infants against RSV during the 2024/2025 season (14,15). Ahead of any mandate from the Department of Health (DoH), the Health Services Executive (HSE) was asked to deliver a limited programme in line with NIAC recommendations (16). In response, the HSE developed a pathfinder programme designed to deliver immunisations, gather evidence of impact, and to identify any enablers and/or barriers to implementation. While a comprehensive evaluation of this programme is now underway, we aimed to provide a rapid analysis of its impact on infant RSV hospitalisations based on immediately available notification data reported to the HSE-Health Protection Surveillance Centre (HPSC).

## Methods

### The RSV immunisation pathfinder programme in Ireland

Infants born from September 1, 2024 to February 28, 2025 were offered nirsevimab in maternity hospitals soon after birth (ideally by the second day), irrespective of their risk for RSV morbidity or mortality. For home births, nirsevimab was offered in the maternity hospital/unit at the next baby check or audiology screening visit (17). From this point we refer to these infants as the *eligible birth cohort*. This is to distinguish them from an additional cohort of high-risk infants under 1 year of age but born before September 1, 2024 who were also targeted for immunisation. High-risk infants in both cohorts included those born before 30 weeks gestation, preterm infants with chronic lung disease, congenital heart disease, pulmonary abnormalities, or neuromuscular conditions impairing upper airway clearance, as well as infants who were expected to be profoundly immunocompromised during the RSV season (14).

### Surveillance data

The data used for this analysis spanned the four previous typical RSV seasons (2018/2020 to 2023/2024, omitting 2020/2021 and 2021/2022 seasons due to the COVID-19 pandemic) and the current 2024/2025 season up until February 28, 2025.

Hospitalised RSV cases were notified to HPSC through the Computerised Infectious Disease Reporting (CIDR) system (https://www.hpsc.ie/CIDR), which is the Irish national database of notifiable infectious disease events, and in accordance with the Infectious Diseases (Amendment) Regulations 2011 (S.I. No. 452 of 2011). For our analysis, notified RSV hospitalisations were aggregated by week within each season, and for the following age groups: <1 year, 1-4 years, 5-14 years, 15-64 years, and those aged 65 years and up. Further, those <1 year of age were split into two groups based on whether their date of birth was before or on/after September 1 (relative to the season), since all those born on or after September 1 during the 2024/2025 season were in the eligible birth cohort.

### Statistical models

To help us understand any potential benefit of nirsevimab immunisation, we fit two generalised additive models (GAM; 16) with negative binomial errors and a log link function to model counts of weekly notified RSV hospitalisations over time, by season and age-group. We first fit a hierarchical GAM (19) with fixed-effects for age-group and season, a smooth (using thin plate splines with 20 knots throughout) for season-week, and additional smooths for season-week by age-group and by season. This *base model* predicts season- and age-group-specific RSV hospitalisation counts across season-weeks, based on their additive effects (on the log scale) while borrowing strength from the marginal smooth for season-week estimated across all age-groups and seasons. To then estimate any benefit of RSV immunisation in the eligible birth cohort, we added to the base model a fixed effect for a binary indicator variable reflecting whether a given week’s count of RSV hospitalisations was observed in the 2024/2025 season *and* in the age-group that would have been eligible to receive nirsevimab based on their date of birth falling on or after September 1, 2024 (the *eligible birth cohort model*). The estimated coefficient for this eligible birth cohort effect was exponentiated to return an incidence rate ratio (and 95% CI). An estimate of averted RSV hospitalisations (and 95% CI) was based on summing the total number of model-predicted weekly hospitalisations in the eligible birth cohort for this season based on this effect estimate, and comparing that to what the model would predict if the effect was equal to zero. Because the base model and the eligible birth cohort model are nested, their respective fits to the data were evaluated using their Akaike information criteria (AIC) and a likelihood ratio test (LRT) of the two models.

All analyses were conducted using the R statistical programming language (version 4.4.1) (20) and the RStudio IDE (version 2024.12.0). GAMs were fit using the mgcv package (18), with additional use of the gratia package for model expiration and interpretation (21). Other key packages included marginaleffects (22), performance (23), sjPlot (24), and ggplot2 (25). The analysis code for running the models and generating predictions and other outputs can be found in the supplemental material; while the code and data will be made available at https://osf.io/6tymf/.

## Results

The observed number of aggregated RSV hospitalisations by age-group and season-week are displayed in Figure 1, along with the corresponding values predicted by the eligible birth cohort model. For each of the previous seasons, RSV hospitalisations were relatively frequent in the eligible birth cohort of infants < 1 year (549 on average, with 655 in 2023/2024), whereas for this season that age-group produced relatively few RSV hospitalisations (154 as of 28/02/2025). Model predictions generally corresponded well with the observed data. A closer look at model fit to the observed data can be found in Figure 2.

**Figure 1.**
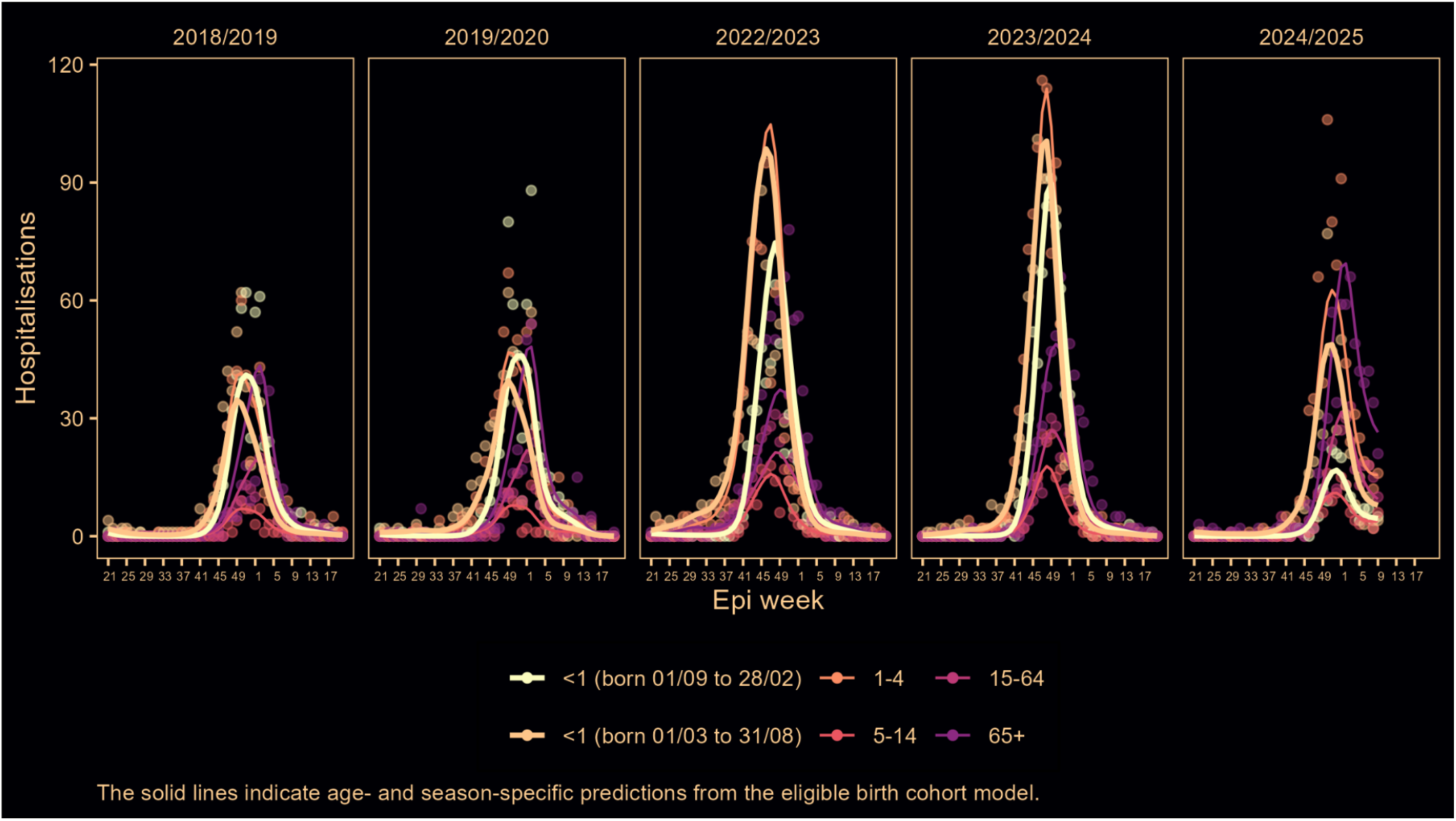
Observed weekly RSV hospitalisations and model predicted values by age-group and season in Ireland

**Figure 2.**
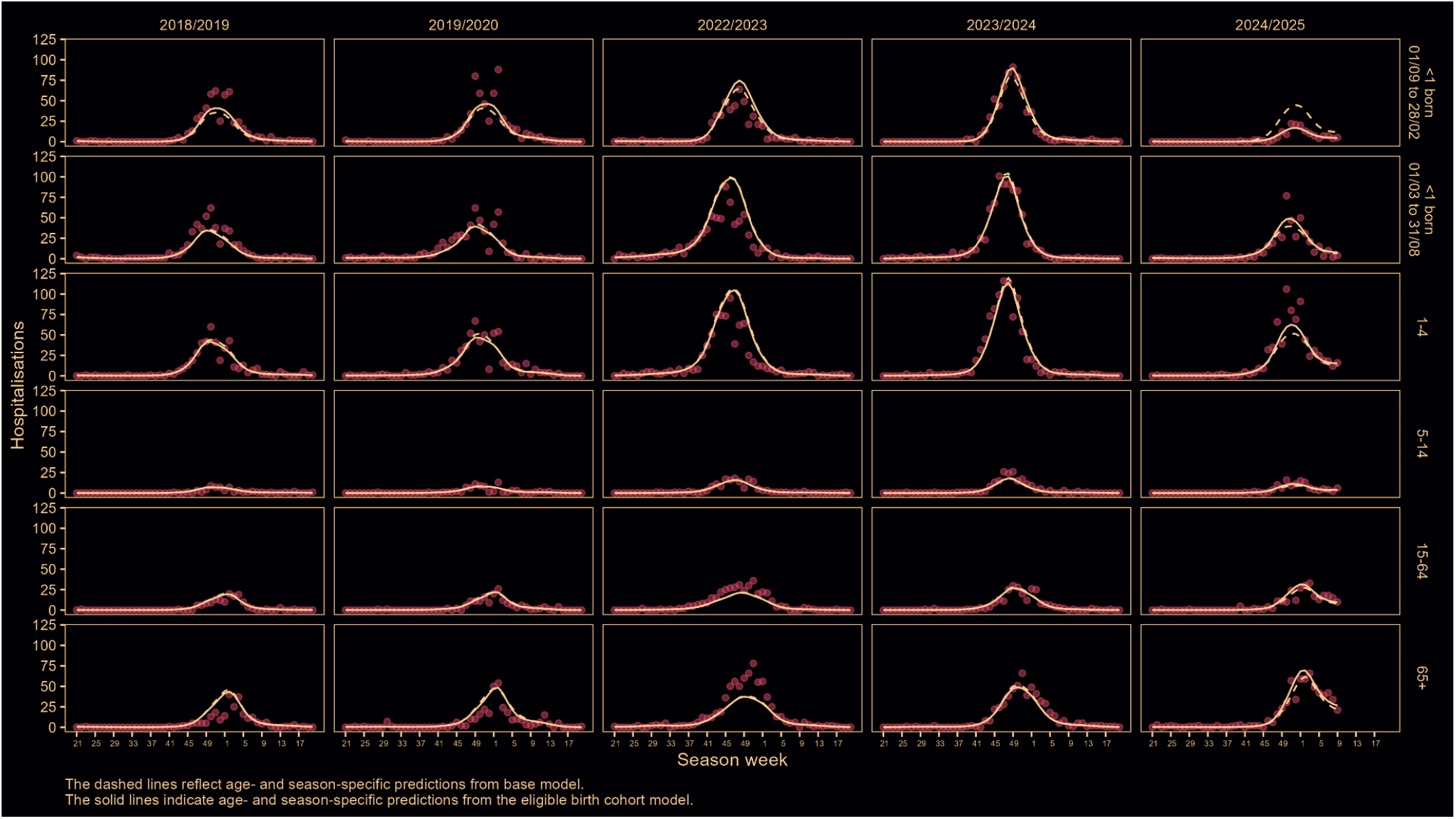
Observed weekly RSV hospitalisations and model predicted values by age-group and season in Ireland, comparing the base and eligible birth cohort models

Figure 3 focuses on observed data in this season’s nirsevimab eligible birth cohort of infants <1 year of age, and compares predictions from the base model to those of the eligible birth cohort model. The latter clearly fits the data better (AICs: 5671.8 vs 5583.9, LRT p-value = < 2.2e-16). The difference between the two curves, each representing the respective models’ predictions, visualises the effect of nirsevimab. Based on the eligible birth cohort model, there was a 75% reduction in the incidence rate for nirsevimab eligible infants born this season, conditional on the age-group and season effects adjusted for in the model (incidence rate ratio 0.25; 95% CI 0.19 to 0.33). This corresponds to an estimated 532 RSV hospitalisations averted (95%CI 369 to 695) for this group, in this season.

**Figure 3.**
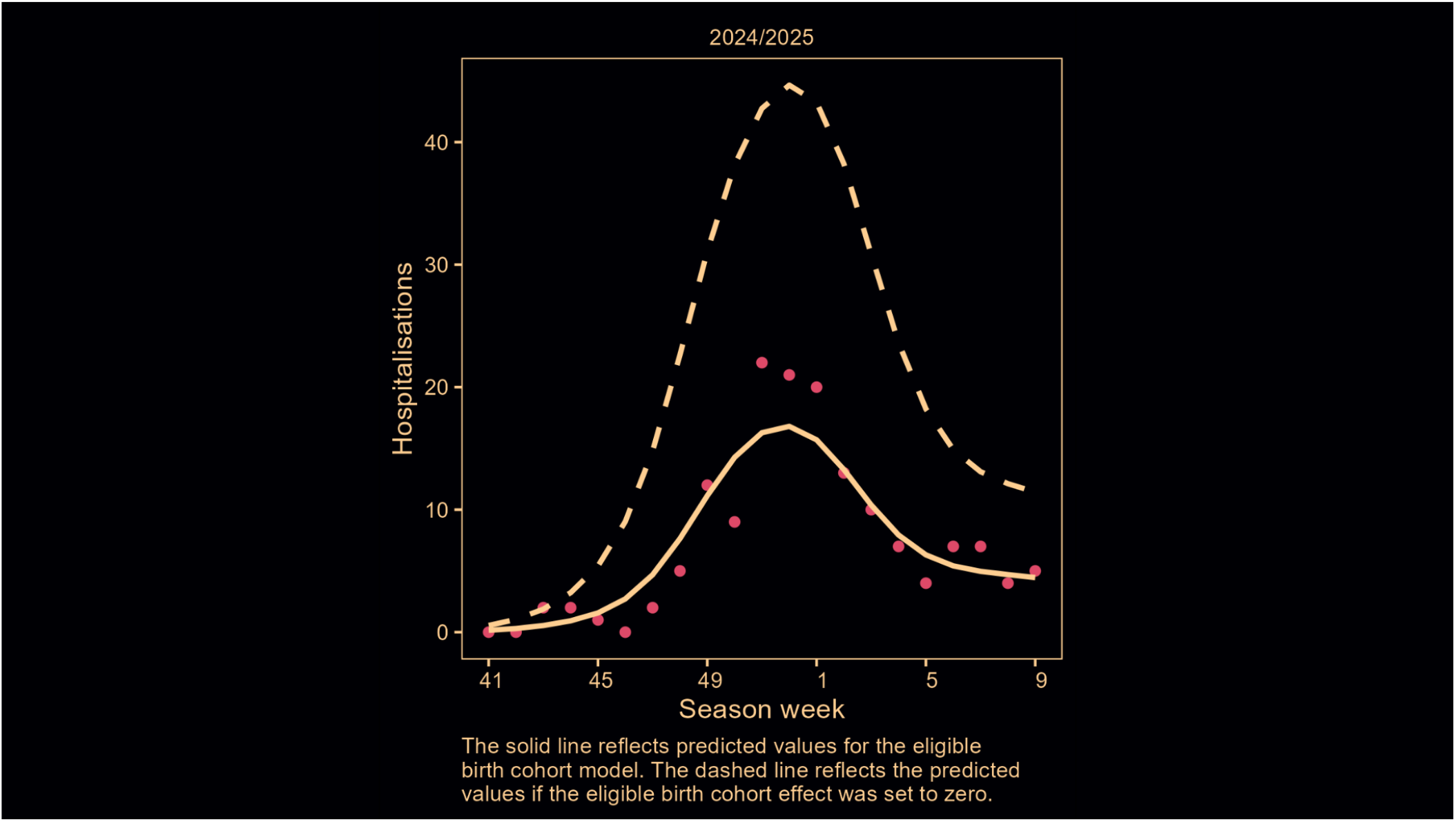
Observed weekly RSV hospitalisations and model predicted values in the eligible birth cohort of infants < 1 year of age for the 2024/2025 season, comparing the base and eligible birth cohort models

## Discussion

RSV presents a significant health risk to young infants and substantially burdens already stretched health systems every winter. Ireland launched a RSV immunisation pathfinder programme for newborns and other clinically vulnerable infants in September of 2024, at the start of the 2024/2025 season (16). While the HSE is conducting a comprehensive evaluation of the Irish programme, we aimed to rapidly analyse the impact of the programme on RSV hospitalisations based on immediately available surveillance notification data.

For our modelling approach, we first estimated a base model to establish an expected age-group and season-specific pattern of RSV hospitalisations over time, based on additive age-group and season specific effects, with no interaction between the two. Despite the relative simplicity of the model, its predictions fit the observed data well, including this season’s data for each age-group, but with the exception of the cohort of infants eligible for the immunisation programme who experienced considerably fewer RSV hospitalisations than this model would suggest. We then built upon this model by adding a binary indicator variable that captured this deviation from the overall pattern established with the base model. This second model was a clear improvement in model fit to data. This best fitting model suggested there was a 75% reduction in infant RSV hospitalisations in the eligible birth cohort this season compared to what we might have expected for this group based on data from other seasons and age groups.

For the purposes of estimating a causal effect of the immunisation programme, our data were inherently limited by their observational nature. However, the base model makes predictions for 30 total groups (six age-groups by five seasons) and one of the few groups to appreciably deviate from this model was the eligible birth cohort for this season, who of course were a key target of the immunisation programme. The data were also limited in that we could not directly confirm whether individual infants were actually immunised, and instead had to rely on their dates of birth as an instrument for immunisation in the aggregate data. This concern is lessened by the high level of programme uptake observed in Ireland (83% overall, and 99% among high risk infants (26)). Finally, our reliance on birth dates as an instrument for immunisation means that any benefits of nirsevimab to infants in the high risk cohort born before September 1, 2024 are not reflected in our estimates, and remain unstudied. Additional, comprehensive analyses accounting for nirsevimab effectiveness, risk-level, and uptake are ongoing.

Despite the limitations of this study, our analysis resulted in estimates of relative benefit that were close to those observed in other studies, including an efficacy of 74.5% (95%CI 49.6 to 87.1) from the MELODY trial (10); an 83.2% reduction in RSV-related infant hospitalisations (95%CI 67.8 to 92.0) estimated in the HARMONIE trial (27); and an evaluation of the Spanish infant nirsevimab programme suggesting that nirsevimab was between 70% and 84% effective in preventing RSV hospitalisations in infants (13). However, because our analysis was based on readily available notification data on aggregated RSV hospitalisations, we were able to provide this evidence of benefit immediately following the peak of the 2024/2025 RSV season, helping to facilitate planning and recommendations for 2025/2026.

## Data Availability

All data produced in the present study are available upon reasonable request to the authors, and will be made publicly available following the final publication of the analysis.

https://osf.io/6tymf/

